# Promoting healthy and sustainable diets through food service interventions in university settings: a scoping review

**DOI:** 10.1101/2024.01.11.24301108

**Authors:** Suzie Kratzer, Melissa A. Theurich, Theresa Mareis, Simone Pröbstl, Nicole Holliday, Sebrina Yan, Anna Leibinger, Ina Monsef, Leonie Bach, Lukas Schwingshackl, Aline Simonetti, Monika Hartmann, Dominic Lemken, Peter von Philipsborn

## Abstract

**Background:** Food service operations in universities and colleges, such as cafeterias and canteens, may support healthy and sustainable diets among students and staff and contribute to a transformation of the wider food system. Multiple studies on interventions to promote health and sustainability in this setting have been conducted, but no up-to-date, comprehensive review exists. This study aims to fill this gap.

**Methods:** We used state-of-the-art scoping review methodology. We included any study examining interventions in university or college food service settings aimed at, or potentially suitable for: 1), supporting healthy and/or sustainable diets, 2) reducing food waste, or 3) otherwise increasing the sustainability of the food service operation (e.g. by improving energy efficiency). We considered studies using any study design published in any language without publication date restrictions. We comprehensively searched six academic databases and conducted forward and backward citation searches. We extracted and charted data on key study characteristics, including the reported direction of effects/associations.

**Results:** We identified 206 studies reporting on 273 interventions. Most studies (71%) used quasi-experimental study designs, were conducted in North America (53%) or Europe (34%), and were implemented in cafeterias or canteens (76%). The most common intervention types were labelling interventions (26%), improved or increased offerings of healthy and/or sustainable food and beverage options (24%), and information and awareness-raising interventions (18%). The most commonly assessed outcomes were implementation-related (e.g. costs, feasibility, acceptability), diet-related (e.g. sales or consumption of specific foods), and sustainability-related (e.g. carbon footprint). Most, but not all, studies reported mostly effects in the desired direction (e.g. increased vegetable consumption, or decreased food waste).

**Discussion:** Multiple approaches for promoting health and sustainability in university and college food service settings exist. The findings of this review suggest that such interventions can be effective, feasible, cost-effective, and aligned with customer and stakeholder expectations. We therefore suggest that they are considered for implementation more widely. Implementation should be accompanied by monitoring and methodologically robust evaluations to allow for evidence-informed tailoring and adjustments and to improve the existing evidence base.

## Background

The global food system faces enormous challenges in terms of human health and environmental sustainability (1). Suboptimal dietary patterns are is among the leading risk factors for chronic disease and premature death worldwide (2, 3). In addition, the global food system is responsible for a quarter to a third of global anthropogenic greenhouse gas emissions, thus contributing substantially to climate change (1, 4). The food system is also the primary driver behind a number of further processes of global environmental change, including biodiversity and habitat loss, the depletion of freshwater resources, deforestation, and land degradation (1). At the same time, approximately one-third of global food production goes to waste either during the production process or at the consumer stage (5). Besides, there are concerns about human trafficking, child labor, and other forms of exploitation in the global food system (6). To safeguard both human and planetary health in the Anthropocene, a shift towards healthier and more sustainable dietary patterns diets is therefore crucial (1). Reducing food waste and a move to more sustainable production practices along the food supply chain are also vital to reduce the environmental impacts of the food system (1).

Institutions of tertiary education (universities, colleges, etc.) can contribute to addressing these challenges in various ways, including through their core functions of education, research, and societal outreach (7, 8). A further area in which universities and colleges can contribute to addressing these challenges is campus food services, such as cafeterias, canteens, kiosks, and cafés (9, 10). Campus food services are generally used by university faculty, staff, and students alike. Interventions in this setting, therefore, hold the potential to positively influence diets of a wide range of audiences. In most institutions, both students and staff spend a substantial part of their time, and often consume a considerable part of their diet, on campus, underlining the importance of these settings. Most students of these institutions are young adults, transitioning from youth to more independent lifestyles in adulthood. This time period has been shown to be a critical phase, during which changes in dietary habits may occur that may last for the rest of adulthood (11). Exposure to canteen foods has also been shown to be associated with dietary patterns outside of the university and college setting (12). University food services may also serve as role models for food services in other settings, such as schools (including high schools) and other workplaces. Besides, exposure to healthy and sustainable diets on campus might shape students’ expectations of food service later in life, e.g. in workplace settings or as parents in school settings. Positive experiences with healthy and sustainable campus food services may, therefore, enable and empower students to act as change agents for a wider transformation of the food system. Interventions to support healthy and sustainable diets in this setting may also have more immediate positive effects. In particular, studies have shown that improving diets among university and college students can have positive effects on their physical and mental health (13-15).

University food services are, therefore, a potentially important setting for interventions to promote health and sustainability. A substantial number of studies have been conducted on such interventions. While there exist several reviews on related topics those are more limited in scope. No comprehensive, up-to-date review of the evidence current research landscape from studies promoting health and sustainability in a university or college setting exists. Therefore, this scoping review aims to identify interventions implemented in university and college food service settings that aim 1) to support healthy and/or sustainable diets, 2) to reduce food waste, or 3) to improve the sustainability of food service operations.

## Methods

### Overview

We used state-of-the-art scoping review methodology as recommended by the Joanna Briggs Institute (JBI) and the PRISMA-ScR reporting guideline (the PRISMA-ScR checklist is shown in Table s1 in the supplementary material) (16, 17). The protocol for this review was registered and published with the Open Science Framework (OSF) at https://doi.org/10.17605/OSF.IO/CM8VA (18) before conducting the literature search.

### Eligibility criteria

We included any studies (except reviews) on interventions implemented in university and college food service settings that aimed to promote healthy and/or sustainable diets, reduce food waste, or otherwise increase the sustainability of food service operations (e.g. through increased energy efficiency). Specifically, studies had to meet the following eligibility criteria:

#### Population

Any person using food services on university or college campuses or connected to such. This includes students, faculty, staff, or visitors, as well as relevant stakeholders like canteen staff or managers, or food service industry suppliers.

#### Intervention

Any intervention aimed at, or potentially suitable for promoting healthy and/or sustainable diets, reducing food waste, or otherwise increasing the environmental sustainability of food service operations. We included studies focusing on either health or environmental sustainability, or both. Examples include:

i. Interventions to increase sales and consumption of healthy and/or sustainable foods (e.g. fruits, vegetables, legumes, whole grains) or decrease sales and consumption of less healthy and/or less sustainable foods (e.g. animal-based foods or ultra-processed foods high in sugar, sodium, and fat).
ii. Interventions to reduce the environmental footprint of foods sold and consumed (e.g. increased offerings of attractive vegetarian meal options, carbon labelling, sourcing local, organic, or seasonal food).
iii. Interventions to reduce food waste.
iv. Interventions to make the food service itself more sustainable (e.g. reduced packaging or other waste, better recycling, higher energy efficiency).

#### Outcomes

Any outcome falling into one or several of the following categories:

i. Diet quality (e.g. food and meal choices, food and nutrient intake of canteen users).
ii. Health outcomes (e.g. body weight, Body Mass Index (BMI), prevalence and incidence of diet-related health outcomes such as obesity or hypertension, mental health outcomes).
iii. Sustainability outcomes (e.g. level of greenhouse gas emissions, adherence to the planetary health diet, level of animal-derived protein intake, level of food waste, water usage).
iv. Implementation-related outcomes (e.g. feasibility, acceptability, costs, personell needs).

#### Comparison

No or alternative interventions or business as usual (where applicable).

#### Setting

Any food service operation in universities, colleges, and similar institutions of tertiary education. This includes canteens, cafeterias, and dining halls, as well as kiosks or vending machines on campus. Studies on privately operated restaurants or cafés located on campuses were not included.

#### Study design

Any study design (including theoretical and conceptual papers) except reviews, commentaries, and letters not reporting on a systematic investigation.

#### Publication type

Any form of academic literature (peer-reviewed or non-peer-reviewed, including preprints) as well as grey literature (i.e. literature that is published in formats other than as academic journal articles, e.g. as government or NGO reports).

#### Language

We did not set any a priori language restrictions, but we conducted the searches in English only. Within our team, we were able to cover studies written in English, French, German, Spanish, and Portuguese.

#### Publication date

No a priori publication date restrictions were set, and databases were searched from their onset.

### Search methods for identification of studies

We systematic searched six academic databases (MEDLINE, the Cochrane Library, Scopus, Epistemonikos, CAB Abstracts, and ERIC) and used references of existing reviews and key primary studies for forward and backward citation searches (i.e. snowballing searches) in Scopus. The search syntax for the database searches was based on three search concepts: i) food service; ii) tertiary education settings; and iii) diet, health, and sustainability. The full search syntax strategies for all databases and the references used for the snowballing searches are shown in the supplementary material. The literature searches were designed and conducted by an information specialist (IM). Searches were conducted on June 8, 2023. We used Endnote for de-duplication.

### Title and abstract screening

We used the web-based application Rayyan for title and abstract screening (19). References were screened by one review author (SK, TM, SP, MT, NH, AL, or AS). Only studies that were clearly irrelevant were excluded, and studies marked as unclear were screened in duplicate by a second review author (SK or PvP). For references identified through MEDLINE, Scopus, and the snowballing searches (constituting 38% of all references), we conducted a full manual screening. For references identified through CAB Abstracts, Epistemonikos, Cochrane Library, and ERIC, we restricted manual screening to references with a high to medium predicted relevance based on Rayyan’s prediction algorithm, excluding references with a star rating below 2.5 out of 5. Rayyan’s prediction algorithm calculates the likelihood that references will be eligible for inclusion based on prior manual screening decisions using artificial intelligence technology. The algorithm has been validated and shown to be reliable (20-22). One validation study found that after screening 20% of all references manually, a star rating of 2.5 or higher had a sensitivity of 100% for identifying eligible studies, meaning that exclusion of references with a star rating below this threshold did not exclude any eligible studies (23).

### Full text screening

Full texts were screened by one review author (SK, TM, AS, SP, NH, LB, or AL), and unclear cases were screened in duplicate by a second review author (SK or PvP). The reasons for excluding studies during the full-text stage were documented.

### Data extraction

Data extraction was done by one review author (SK, NH, SP, or TM) using a data extraction form in Google Sheets shown in the supplementary material. Any uncertainties were marked by the reviewer and double-checked by a second review author (SK or PvP). We extracted data on study design, setting, intervention characteristics, outcomes, and the reported direction of effect.

### Data synthesis

We summarized results narratively, in tables, and graphically in an Evidence Gap Map. An Evidence Gap Map is a visual tool designed to provide an overview of the existing evidence on a topic, highlighting gaps in the evidence base while also showing where evidence is more abundant. We used the NOURISHING Framework, developed by the World Cancer Research Fund International, to classify interventions (24).

### Quality assurance measures

We implemented several quality assurance measures. Among others, we piloted the procedures at each stage of the review process, conducted regular team meetings, drafted internal guidance documents, and kept a list of rolling questions that were updated throughout the review process to ensure clarity and consistency between review authors. As a calibration exercise, review authors involved in the screening (SK, TM, AL, NH, SP, AS, and PvP) independently assessed a set of references (50 at title and abstract stage, and 10 at full-text screening stage). Following this, inconsistencies and any questions that arose were discussed in the team and the guidance documents were adjusted accordingly. The data extraction sheet was pilot-tested by two review authors (SK, PvP) on five studies and adjustments were made where needed.

### Results

### Results of the search and selection process

The database and snowballing searches yielded a total of 22,461 records before de-duplication and 17,815 after de-duplication. Of these, 6,917 records were identified through MEDLINE, Scopus, and the snowballing searches, and were manually screened in full. We then applied Rayyan’s prediction algorithm to the remaining 10,898 records, excluding 9,730 records that received a star rating of below 2.5, leaving another 1,360 records with a star rating of 2.5 or higher to be screened manually. In total, we excluded 17,333 records at title and abstract screening stage, leaving 482 records for full-text screening, of which we included 204 records reporting on 206 studies in our review. Among the studies excluded at the full-text screening stage were 5 studies in Korean and Chinese, for which we were unable to arrange translation. An exemplary list of studies excluded at this stage with reasons for exclusion is provided in table s2 in the supplementary material. The search and screening process is depicted in the PRISMA flow diagram in Figure 1.

**Figure 1.**
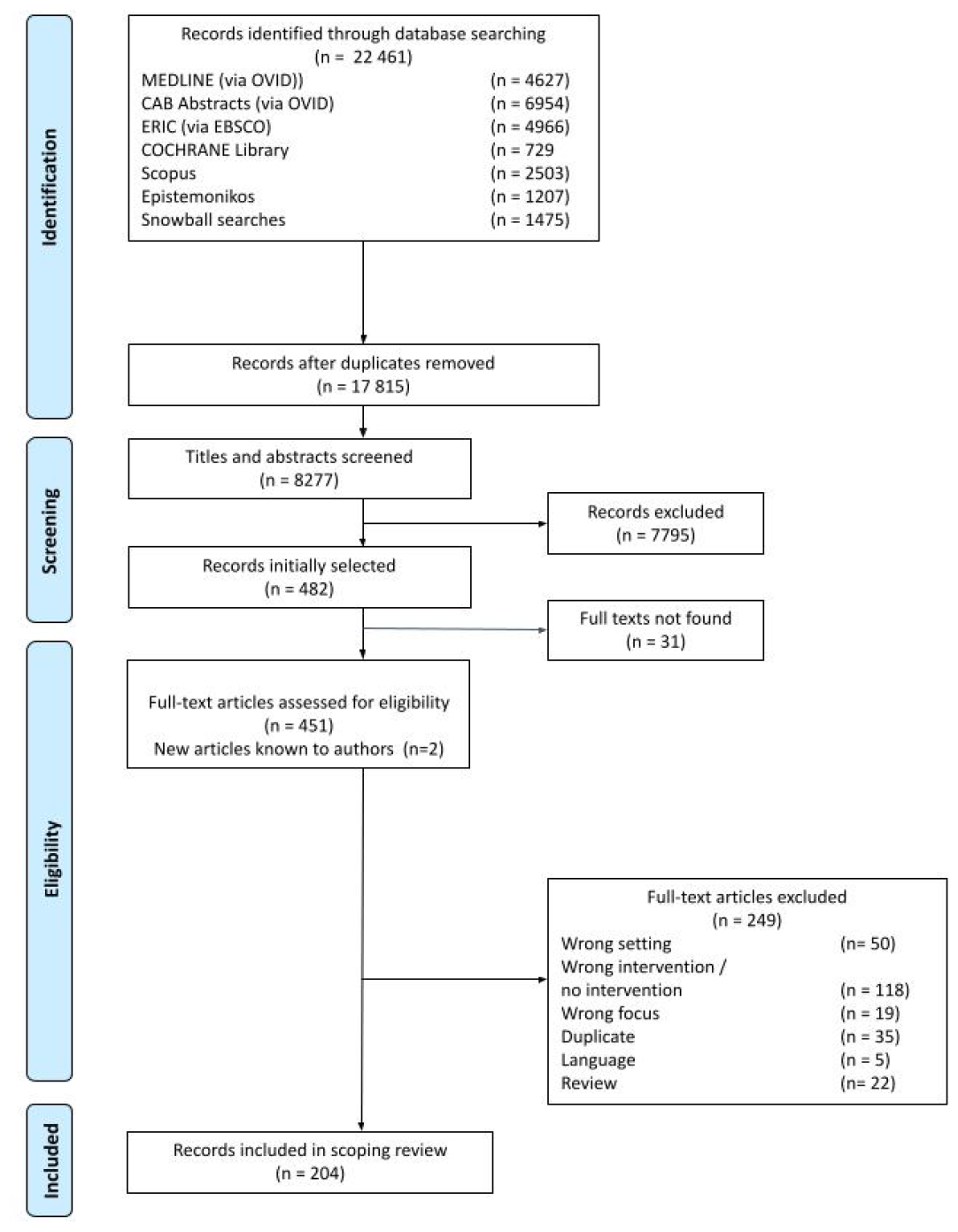
PRISMA Flow Chart

### Characteristics of included studies

A full list of included studies with key study characteristics is provided in the supplementary material. An interactive spreadsheet allowing for filtering by study characteristics is available online at https://osf.io/ykv3n. In the following, we present a short overview.

#### Study design, setting, and country focus

The most common study design was quasi-experimental (including simple before-and-after-studies, and more sophisticated designs such as interrupted time series studies): 146 studies (71% of the total) fell into this category, followed by cross-sectional designs (12%), mixed-methods designs (10%) and qualitative studies (4%). The majority of studies were from North America (53%) or Europe (34%), and only few (13%) from other regions. Regarding countries, most studies were conducted in the US (96 studies), the UK (16 studies), as well as Canada and Germany (13 studies each). With regard to the type of food service operation, three out of four studies focused on cafeterias or canteens (76%), while 10% focused on vending machines on campus, and 7% reported on campus-wide interventions (see Table 1).

**Table 1:**
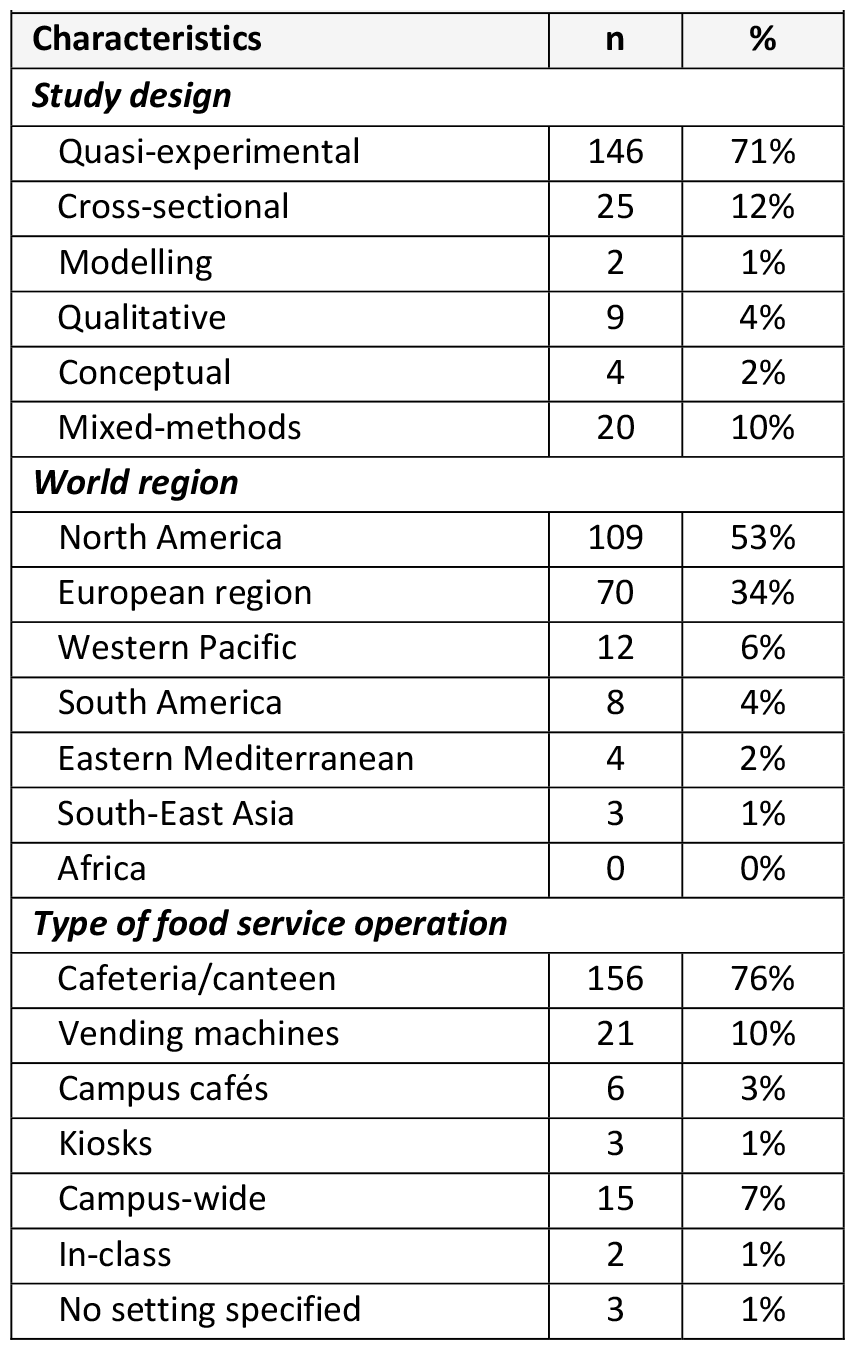
Distribution of study designs, settings, and countries across all included studies.

#### Study focus

We classified the focus of the intervention for the majority of studies (59%, n=121) to be health-related, for 68 studies (33%) to be sustainability-related, and for 17 studies (8%) to cover both (i.e. health and sustainability). As expected, sustainability has only come to the forefront in recent years. Until 2015, there were only 1 to 3 studies published each year with a sustainability focus. For the years after 2015, we found a minimum of 7 studies with a sustainability focus in each year, the maximum (n=18) being published in 2022.

#### Study objective

Most studies (n=128, 62%) were aimed at promoting healthy diets patterns among food service users, while 35 (17%) were aimed at promoting sustainable diets, 28 (14%) aimed at food waste reduction, and 20 (10%) at other approaches for increasing the sustainability of the food service operation. On a more granular level, interventions to promote fruit and vegetable intake (n=21, 10%), to reduce consumption of animal-based foods (n=22, 11%), and to reduce post-consumer food waste (n=22, 11%) were most common (see Table 2).

**Table 2:**
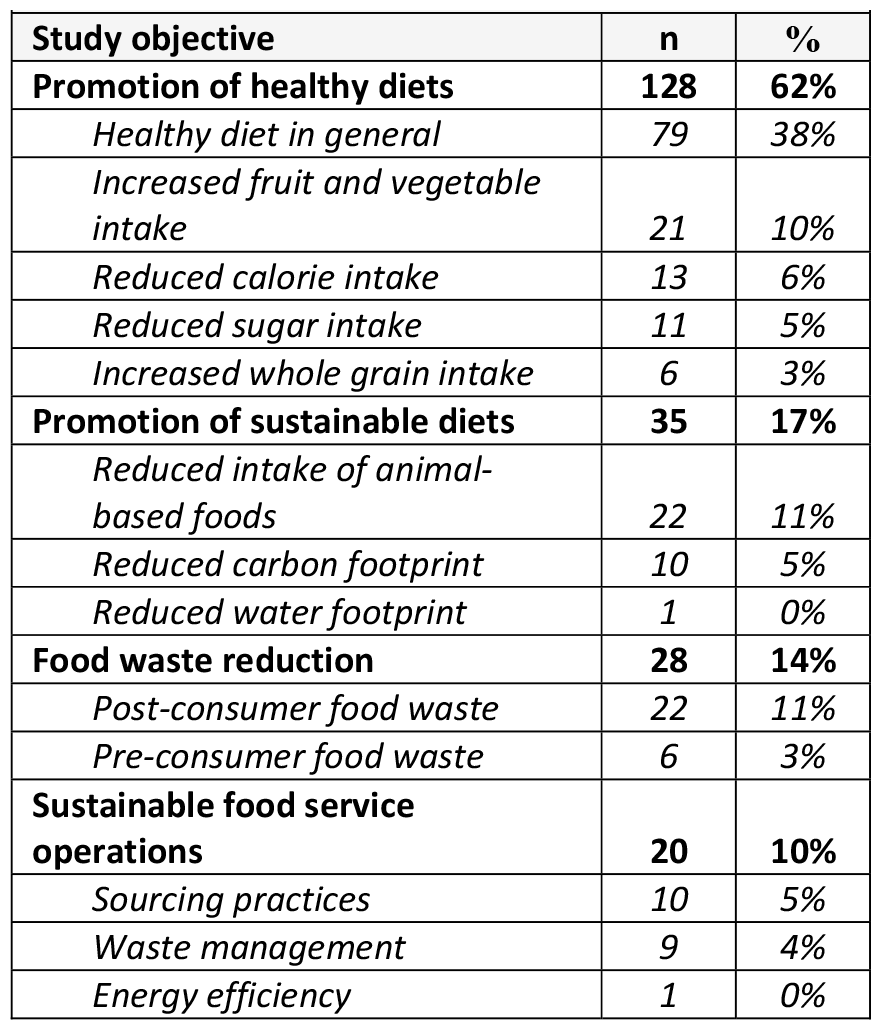
Distribution of study objectives across included studies.

| **Table 2: Distribution of study objectives across included studies** |  |  |
| --- | --- | --- |
| **Study objective** | **n** | **%** |
| **Promotion of healthy diets** | **128** | **62%** |
| *Healthy diet in general* | 79 | 38% |
| *Increased fruit and vegetable intake* | 21 | 10% |
| *Reduced calorie intake* | 13 | 6% |
| *Reduced sugar intake* | 11 | 5% |
| *Increased whole grain intake* | 6 | 3% |
| **Promotion of sustainable diets** | **35** | **17%** |
| *Reduced intake of animal-based foods* | 22 | 11% |
| *Reduced carbon footprint* | 10 | 5% |
| *Reduced water footprint* | 1 | 0% |
| **Food waste reduction** | **28** | **14%** |
| *Post-consumer food waste* | 22 | 11% |
| *Pre-consumer food waste* | 6 | 3% |
| **Sustainable food service operations** | **20** | **10%** |
| *Sourcing practices* | 10 | 5% |
| *Waste management* | 9 | 4% |
| *Energy efficiency* | 1 | 0% |

#### Classification according to the NOURISHING Framework

In total, 273 distinct interventions were examined in the 206 included studies (some studies reported on more than one intervention). Of those, 209 aimed at changing the food environment, 7 at changing the food supply chain, and 64 applied behaviour change communication techniques. The most common intervention types were labelling interventions (n=71), increased or improved offerings of healthy and sustainable options (n=65), and information and awareness raising interventions (n=50) (see Table 3).

**Table 3:** Classification of included studies according to intervention type.

|  |  |  |
| --- | --- | --- |
| **Intervention area (based on the NOURISHING framework)** | **n (%)** | **Examples** |
| **Food environment interventions** |  |  |
| Labelling interventions | 71 (26%) | Health-related labelling: nutritional traffic light labels, calorie labelling, health score labelling, nutritional values shown next to meals.<br>Sustainability-related labelling: carbon / CO<sub>2</sub> labelling, labelling with sustainability ratings. |
| Offer healthy and sustainable food | 65 (24%) | Changing the offer towards a greater choice of healthier or more sustainable options (e.g. more healthier snack options in a vending machine, less sugar-sweetened beverages on offer, offering more vegetable side dishes in the cafeteria).<br>Nudging interventions directing the consumers towards healthier or more sustainable choices (e.g. changing the order of meals, making certain foods more salient). |
| Economic tools | 15 (5%) | Price reduction for healthier or more sustainable options (e.g. the vegetarian option, salad instead of fries, low sugar drinks).<br>Price increases for less healthy or less sustainable options.<br>Rewards for avoiding food waste. |
| Advertisement regulation | 0 | n.a. |
| Improve food quality or sustainability through reformulation | 27 (10%) | Changing recipes to arrive at a better nutrient profile, replacing animal-based proteins with plant-based proteins, using less salt or more whole wheat flour. |
| Other food service interventions | 31 (11%) | Changing the environment to reduce food waste or to promote recycling, changes towards less plastic in the food service. |
| **Food system interventions** |  |  |
| Food supply chain interventions | 7 (3%) | Changing the sourcing of food toward local or organic produce. |
| **Behaviour change communication interventions** |  |  |
| Information and awareness raising interventions | 50 (18%) | Posters and information campaigns on healthy eating, food waste avoidance, correct recycling, and the links between food and sustainability in general. |
| Counselling interventions | 0 (0%) | n.a. |
| Education and skill-building interventions | 7 (3%) | Courses for food service users on how to eat healthily and/or sustainably, training for kitchen staff on how to prepare satisfying plant-based dishes or on how to avoid food waste while preparing meals. |

#### Length of follow-up

The length of follow-up differed considerably between studies. Most studies were of short duration, with 57% lasting less than a month. In 43 studies (28%), the follow-up lasted between one and six months; in three studies (2%), between 6 and 12 months; in 17 studies (11%), follow-up was longer than one year; and two studies did not report the length of follow-up. In 54 studies, length of follow-up was not applicable due to the study design (e.g. cross-sectional design, or formative research reporting on proposed or potential interventions).

#### Outcomes

The most commonly reported types of outcomes were implementation-related outcomes, including acceptability, feasibility, implementation fidelity, uptake, and views on (potential) interventions (n=107). This was followed by outcomes related to diet quality, e.g. changes in sales or intake of specific food items, as well as changes in energy or nutrient intake (n=104). The third largest group were sustainability outcomes (n=76), including changes towards more plant-based diets or reduction of food waste. Economic outcomes, like costs or profitability; mental health-related outcomes, like weight concerns; and diet-related health outcomes, like cardiovascular risk factors or over-/under- weight, were examined in much smaller numbers (11, 6, and 1, respectively).

### Evidence Gap Map

The Evidence Gap Map (see Figure 2) provides a visual summary of the interventions and outcomes explored in the included studies, along with the corresponding methods used to investigate each pairing of intervention and outcome. Each pair is also marked with a code indicating the respective study, as listed in the list of included studies provided online at https://osf.io/ykv3n. Codes starting with A (e.g. A1, A2, etc.).) indicate studies aimed at supporting healthy and sustainable dietary patterns; B stands for studies aiming to avoid food waste; and C for studies aiming at otherwise improving the sustainability of food service operations. By far, the largest group of intervention-outcome-pairs are represented by diet quality outcomes for ‘Labelling interventions’ and for ‘Offer healthy and sustainable food’, with 42 outcomes of diet quality measured for each. ‘Acceptability’ was also studied across all intervention types, mostly for ‘Labelling interventions’ (17) and for ‘Information and awareness raising interventions’ (16). Most other intervention-outcome combinations are somewhat populated in the table, however, there are some notable gaps. ‘Labelling interventions’ were the only interventions that had been evaluated for mental health outcomes. Also, only one health-related outcome was reported for ‘Offer healthy and sustainable food’ and ‘Information and awareness raising interventions’, notably from the same study.

**Figure 2.**
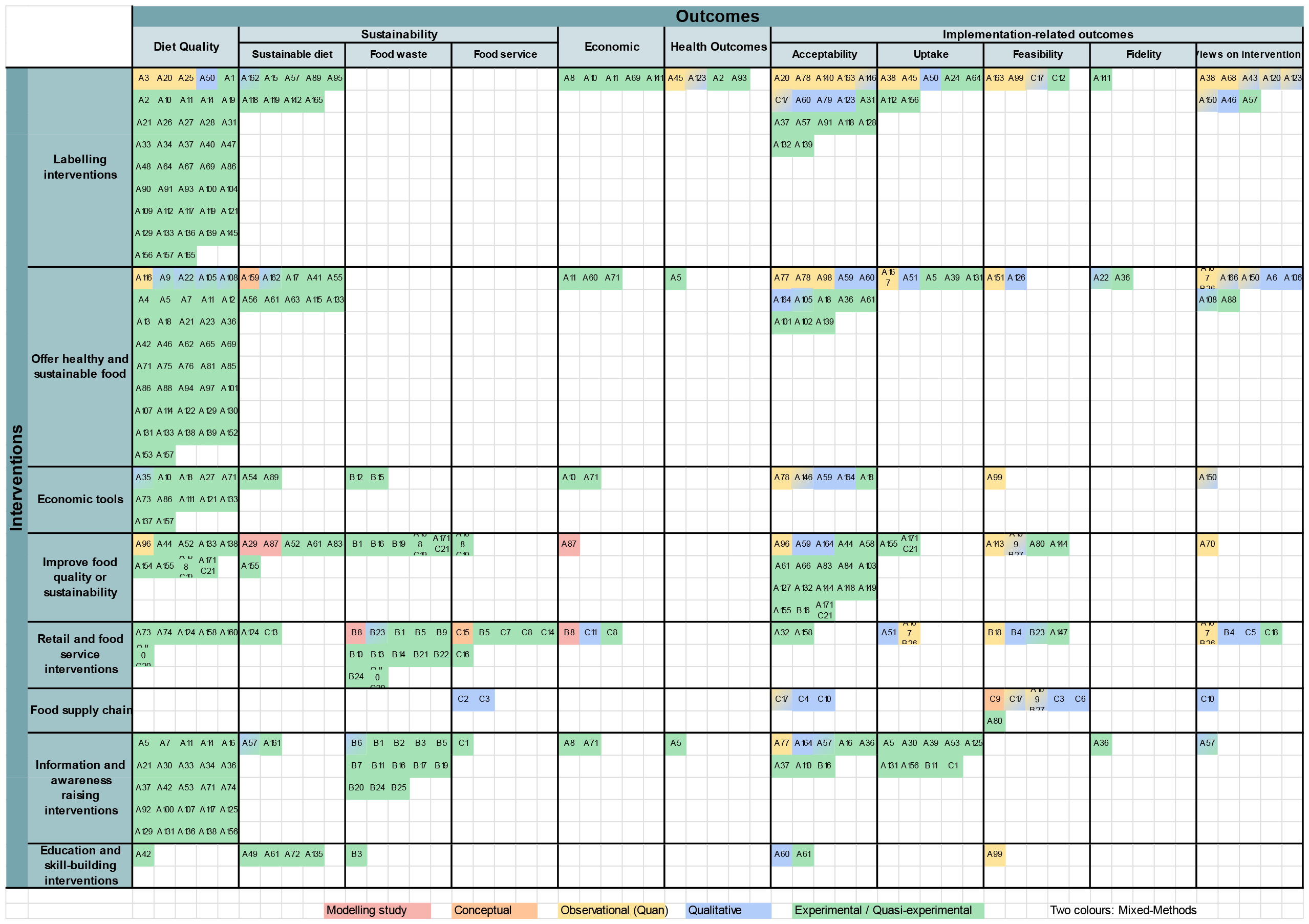
The Evidence Gap Map.

### Effects of interventions

Of those studies that reported effects of interventions, most reported positive beneficial effects, i.e. effects in the desired direction (e.g. increased vegetable intake, reduced food waste) (see supplementary material). We extracted information on the reported direction of the effect but did not assess effect sizes, nor did we conduct a quality appraisal. Findings on the effects of interventions should, therefore, be interpreted with caution.

## Discussion

### Summary of main findings

A large number and variety of approaches to promote health and sustainability in university and college food service settings exist. These include, among others, health and sustainability labels; increased and improved offerings of healthy and sustainable options; nudging interventions to facilitate healthier and more sustainable choices; economic instruments such as price increases or rebates; and a variety of behaviour change communication measures. We identified 206 studies examining such interventions. The most common study designs were quasi-experimental, and cross-sectional designs and qualitative studies. The majority of studies were conducted in a small number of countries, in particular the US, Canada, the UK, and Germany. Most studies focused on health, and only a limited number considered both health and sustainability explicitly. Implementation- and diet-related outcomes were most frequently assessed. Most, but not all, studies reported effects in the desired direction, such as increased intake of fruit and vegetables or a decrease in food waste.

### Strengths and limitations

Our review has a number of important strengths. We used state-of-the-art scoping review methodology as recommended by the JBI Handbook and the PRISMA-ScR reporting guideline, which are considered methodological gold standards in the field (16, 17). Our review is based on an a-priori protocol which we prospectively registered before searches were conducted (18), and all differences between protocol and review are explained in the supplementary material. The review is based on comprehensive literature searches in six databases conducted by an experienced information specialist. We also implemented several quality assurance procedures to ensure consistency among review authors and to minimizeminimise the risk of errors. The types of interventions synthesised in this scoping review include a broad range of settings within the university and college food services, including cafeterias, canteens, cafes, and vending machines.

Our review also has a number of limitations. Screening and data extraction were not done fully in duplicate. We did not screen all references manually, but partly relied on an artificial intelligence-based prediction algorithm, which has not yet been validated for food environment and systems research. Data extraction was done by one review author only, and only double-checked by a second review author when the first indicated uncertainties. We assessed only the reported direction of effects and not effect sizes, and we did not conduct a quality appraisal of included studies, and did not assess the outcome-specific certainty of evidence. We also recognize some difficulties in the terminology around campus food service operations across countries. While we excluded studies that were clearly about privately-operated restaurants or cafes that were located on or around campus that were not otherwise connected to a university or college, we acknowledge that the distinction between such private establishments and university food service operations may not always be clear-cut.

### Implications for research

A number of implications for research arise from the findings of our review. This pertains, among others, to the types of outcomes examined. The number of studies examining both health and sustainability outcomes is growing but is still limited. Given the importance of diets and the broader food system to both human and planetary health, the joint examination of both health and sustainability outcomes should be routinely considered in future studies on the topic. Implementation- related outcomes (such as costs, acceptability, perceived feasibility, etc.) were examined in a substantial number of studies. Given the fact that these aspects can be crucial for sustainability and the scaling-up of interventions, these outcomes should be maintained in future investigations. Mental health outcomes, such as the effects on symptoms of eating and body image disorders (e.g. orthorexia and anorexia) were only examined in a limited number of labelling studies. Given the high prevalence of these disorders and related symptoms, particularly among young adults, and the plausibility of effects of diet-related interventions on them, this is a striking gap, which should be addressed in future research.

Other methodological aspects also have implications for research. Most studies were of short duration, with a follow-up of less than a month. Given the fact that effects may change over time – accumulating or petering out – and that implementation-related factors may equally depend on the implementation phase, studies with longer follow-up periods would be desirable. Most studies used relatively simpleless trustworthy study designs (the most common were uncontrolled before-after studies). While randomized controlled trials (RCTs) may in many cases not be feasible, more sophisticated quasi- experimental methods such as interrupted time series (ITS) studies may be and should be used more often. Most of the studies in our review that reported on diet-related outcomes relied on aggregated data, most typically sales data not linked to individual customers. While such data can provide a reliable measure for average effects, they do not allow for analyses stratified by customer group, e.g. vegetarians versus non-vegetarians. One of the few studies doing so showed that a price reduction on vegetarian dishes and a price increase on meat dishes mostly changed the consumption behaviour of those who already ate very little meat before the intervention and had little effect on those in the “least vegetarian” quartile, i.e. those who ordered few vegetarian or vegan meals before (25). This shows that a more detailed view on the effects of such interventions is warranted and should be included in future research in order to be able to design interventions better addressing certain target groups.

### Implications for policy and practice

A large variety of approaches for promoting health and sustainability in the setting of university and college food services, such as cafeterias and canteens, exist. These include, among others, health and sustainability labels; increased and improved offerings of healthy and sustainable options; nudging interventions to facilitate healthier and more sustainable choices; economic instruments such as price increases or rebates; and a variety of behaviour change communication measures. Findings of this review suggest that such approaches can be effective in achieving their desired goals of positively impacting health- and sustainability-related outcomes, such as an increased consumption of healthy and sustainable foods, or a reduction in food waste. Results also suggest that the implementation of such interventions can be feasible, cost-effective, and in line with customer and stakeholder expectations.

However, there is also a considerable variation in the observed effects. This may partly be explained by methodological factors (e.g. studies based on small sample sizes with a high baseline variation in outcomes, and short study durations). On the other hand, this may also suggest that factors such as intervention design and intensity, co-interventions, and other contextual factors may influence the effectiveness of interventions. Tailoring interventions to local circumstances as well as monitoring and evaluation to allow for adjustments are, therefore, crucial. While the evidence base for the effectiveness of suitable interventions is growing, important research gaps remain, as explained above. Implementation should ideally be accompanied by methodologically-sound evaluations. For this purpose, food service practitioners and university administrators might best collaborate with researchers in the field. Thorough process evaluation can also be helpful when comparing the results of different studies and therefore aid understanding of which interventions work best in these or similar settings (26)

### Comparison with similar studies

A number of reviews on related topics have been published to date. A scoping review of food and nutrition interventions in post-secondary educational facilities published in 2021 identified 38 publications through a narrower search in three databases restricted to articles published in English between 2015 and 2019 (9). Similar to our review, they concluded that such interventions hold promise to positively impact health and sustainability, but that the existing evidence base should be improved with methodologically more robust evaluations that consider both human and planetary health. In contrast to our review, they did not identify any study examining both health and sustainability outcomes, and only a minimal number of studies on sustainability (9). Indeed, a substantial part of the studies with such a focus included in our review were published after 2019, highlighting the evolution of the field towards a more integrated approach. A recent systematic review of interventions to improve sustainability in food service operations (decreasing food waste or single use-item and packaging waste, or promoting sustainable diets) equally showed significant pro-environmental changes in consumer expectations, behaviours, as well as cognitive and affective attitudes (27).

## Conclusions

Effective, feasible, and economically viable interventions to promote health and sustainability in university and college food services that are aligned with customer and stakeholder expectations exist. In light of this evidence, and the pressing challenges faced by the global food system, we suggest that such interventions should be implemented more broadly while being accompanied by methodologically-sound evaluations to further improve the evidence base.

## Data Availability

All data described in the manuscript is published alongside the article as supplementary material or is available on the Open Science Framework at https://osf.io/ykv3n.

## Declarations

## Acknowledgements

PvP, DL, MH designed research; SK, TM, SP, SY, MT, NH, AL, IM, LB, AS conducted research; SK and PvP analyzed data; PvP, SK and MT wrote the original draft, SK, PvP, MT, TM, SP, NH, SY, AL, IM, LB, LS, AS, MH and DL edited the content. PvP supervised the research. MH, DL and PvP acquired the funding. All authors read and approved the final manuscript.

