## Supplementary material for "Promoting healthy and sustainable diets through food service interventions in university settings: a scoping review"

**Version**: V5, 11.01.2024

### 1. PRISMA ScR Checklist

Preferred Reporting Items for Systematic reviews and Meta-Analyses extension for Scoping Reviews (PRISMA-ScR) Checklist.

| Table s1: PRISMA-ScR Checklist | | | |
|---|---|---|---|
| **Section** | **Item #** | **PRISMA-ScR Checklist item** | **Reported on page** |
| **TITLE** | | | |
| Title | 1 | Identify the report as a scoping review. | 1 |
| **ABSTRACT** | | | |
| Structured summary | 2 | Provide a structured summary that includes (as applicable): background, objectives, eligibility criteria, sources of evidence, charting methods, results, and conclusions that relate to the review questions and objectives. | 2 |
| **INTRODUCTION** | | | |
| Rationale | 3 | Describe the rationale for the review in the context of what is already known. Explain why the review questions/objectives lend themselves to a scoping review approach. | 3-4 |
| Objectives | 4 | Provide an explicit statement of the questions and objectives being addressed with reference to their key elements (e.g., population or participants, concepts, and context) or other relevant key elements used to conceptualize the review questions and/or objectives. | 4 |
| **METHODS** | | | |
| Protocol and registration | 5 | Indicate whether a review protocol exists; state if and where it can be accessed (e.g., a Web address); and if available, provide registration information, including the registration number. | 4 |
| Eligibility criteria | 6 | Specify characteristics of the sources of evidence used as eligibility criteria (e.g., years considered, language, and publication status), and provide a rationale. | 4-5 |
| Information sources* | 7 | Describe all information sources in the search (e.g., databases with dates of coverage and contact with authors to identify additional sources), as well as the date the most recent search was executed. | 5 |
| Search | 8 | Present the full electronic search strategy for at least 1 database, including any limits used, such that it could be repeated. | Supplementary Material |
| Selection of sources of evidence† | 9 | State the process for selecting sources of evidence (i.e., screening and eligibility) included in the scoping review. | 5-6 |
| Data charting process‡ | 10 | Describe the methods of charting data from the included sources of evidence (e.g., calibrated forms or forms that have been tested by the team before their use, and whether data charting was done | 6 |

| Table s1: PRISMA-ScR Checklist | | | |
|---|---|---|---|
| **Section** | **Item #** | **PRISMA-ScR Checklist item** | **Reported on page** |
| | | independently or in duplicate) and any processes for obtaining and confirming data from investigators. | |
| Data items | 11 | List and define all variables for which data were sought and any assumptions and simplifications made. | 6, Supplementary material |
| Critical appraisal of individual sources of evidence§ | 12 | If done, provide a rationale for conducting a critical appraisal of included sources of evidence; describe the methods used and how this information was used in any data synthesis (if appropriate). | n.a. |
| Synthesis of results | 13 | Describe the methods of handling and summarizing the data that were charted. | 6 |
| **RESULTS** | | | |
| Selection of sources of evidence | 14 | Give numbers of sources of evidence screened, assessed for eligibility, and included in the review, with reasons for exclusions at each stage, ideally using a flow diagram. | 7 |
| Characteristics of sources of evidence | 15 | For each source of evidence, present characteristics for which data were charted and provide the citations. | Characteristics of included studies Tables A, B, C |
| Critical appraisal within sources of evidence | 16 | If done, present data on critical appraisal of included sources of evidence (see item 12). | n.a. |
| Results of individual sources of evidence | 17 | For each included source of evidence, present the relevant data that were charted that relate to the review questions and objectives. | n.a. |
| Synthesis of results | 18 | Summarize and/or present the charting results as they relate to the review questions and objectives. | 8-12 |
| **DISCUSSION** | | | |
| Summary of evidence | 19 | Summarize the main results (including an overview of concepts, themes, and types of evidence available), link to the review questions and objectives, and consider the relevance to key groups. | 13-15 |
| Limitations | 20 | Discuss the limitations of the scoping review process. | 13 |
| Conclusions | 21 | Provide a general interpretation of the results with respect to the review questions and objectives, as well as potential implications and/or next steps. | 15 |
| **FUNDING** | | | |
| Funding | 22 | Describe sources of funding for the included sources of evidence, as well as sources of funding for the scoping review. Describe the role of the funders of the scoping review. | 16 |

#### 2. Search Syntax

##### 2.1. MEDLINE (through Ovid)

\# Searches
1 food services/ or menu planning/ or restaurants/
2 ((food? or meal?) adj2 (service* or system* or purchas* or competitiv*)).tw,kf.
3 (dining* or cafeteria* or restaurant* or canteen* or kitchen? or cuisine? or "out of home" or cater* or vending or kiosk* or snack bar* or snackbar* or cafe* or lunch room* or lunchroom* or foodservice*).tw,kf.
4 or/1-3
5 Students/ or student*.tw,kf.
6 (universit* or "higher education" or college* or post-secondar* or postsecondar* or campus* or institution* or facult* or graduate or undergraduate or postgraduate or bachelor program* or master program*).tw,kf.
7 or/5-6
8 exp "diet, food, and nutrition"/
9 (diet* or eating or nutrition* or meal* or food or foods or lunch or beverage* or drink* or refreshment* or water*).ti,ab,kf.
10 Sustainable Development/ or "Conservation of Natural Resources"/
11 climate/ or climate change/ or global warming/ or Greenhouse Effect/ or Greenhouse Gases/ or carbon footprint/
12 Whole Grains/ or Fruit/ or Vegetables/ or exp Meat/ or exp Vegetarians/
13 (sustainab* or climate or waste* or "eat lancet" or environmental* or carbon or emission? or greenhouse gas* or GHG or global warming or biodiversit* or bio-diversit* or pollution* or planet* or food loss or left over* or leftover* or trash or single-use or reusable or re-usable or locally produced or locally sourced or seasonal or eco* or whole grain* or whole meal? or wholemeal* or whole wheat or refined grain* or fruit* or vegetable* or legume* or fish* or seafood* or fibre? or sugar* or fat* or dairy or sodium or salt* or water* or calorie* or kilocalorie* or meat* or plant based or foodscape* or vegetarian* or vegan* or flexitarian* or health* or unhealth* or wellbeing or well being or balanced or nutritious or obesit* or over weight* or overweight* or body weight or bodyweigth* or BMI or diabetes or NCD* or menu*).tw,kf.
14 or/8-13
15 4 and 7 and 14

##### 2.2. CAB Abstracts

\# Searches
1 exp catering/ or exp dining facilities/ or menu planning/
2 ((food? or meal?) adj2 (service* or system* or purchas* or competitiv*)).ti,ab.
3 (dining* or cafeteria* or restaurant* or canteen* or kitchen? or cuisine? or "out of home" or cater* or vending or kiosk* or "snack bar*" or snackbar* or cafe* or "lunch room*" or lunchroom* or foodservice*).ti,ab.
4 or/1-3

5 college students/ or university students/
6 (student* or universit* or "higher education" or college* or post-secondar* or postsecondar* or campus* or institution* or facult* or graduate or undergraduate or postgraduate or bachelor program* or master program*).ti,ab.
7 or/5-6
8 (diet* or eating or nutrition* or meal* or food or foods or lunch or beverage* or drink* or refreshment* or water*).ti,ab.
9 (sustainab* or climate or waste* or "eat lancet" or environmental* or carbon or emission? or greenhouse gas* or GHG or global warming or biodiversit* or bio-diversit* or pollution* or planet* or food loss or left over* or leftover* or trash or single-use or reusable or locally produced or locally sourced or seasonal or eco* or whole grain* or whole wheat* or whole meal? or wholemeal* or refined grain* or fruit* or vegetable* or legume* or fish* or seafood* or fibre? or sugar* or fat* or dairy or sodium or salt* or water* or calorie* or kilocalorie* or meat* or plant based or foodscape* or vegetarian* or vegan* or flexitarian* or health* or unhealth* or wellbeing or well being or balanced or nutritious or obesit* or over weight* overweight* or body weight or bodyweigth* or BMI or diabetes or NCD* or menu*).ti,ab.
10 or/8-9
11 4 and 7 and 10

#### 2.3. ERIC

### Query
S6 S3 AND S4 AND S5
S5 diet* OR eating OR nutrition* OR meal* OR food OR foods OR lunch OR beverage* OR drink* OR refreshment* OR water* OR sustainab* OR climate OR waste* OR "eat lancet" OR environmental* OR carbon OR emission OR emissions OR "greenhouse gas*" OR ghg OR "global warming" OR biodiversit* OR bio-diversit* OR pollution* OR planet* OR "food loss" OR "left over*" OR leftover* OR trash OR single-use OR reusable OR "re-usable" OR "locally produced" OR "locally sourced" OR seasonal OR eco* OR "whole grain*" OR "whole wheat" OR "whole meal*" OR wholemeal* OR "refined grain*" OR fruit* OR vegetable* OR legume* OR fish* OR seafood* OR fibre OR fibres OR sugar* OR fat* OR dairy OR sodium OR salt* OR water* OR calorie* OR kilocalorie* OR meat* OR "plant based" OR foodscape* OR vegetarian* OR vegan* OR flexitarian* OR health* OR unhealth* OR wellbeing OR "well being" OR balanced OR nutritious OR obesit* OR "over weight*" OR overweight* OR "body weight*" OR bodyweigth* OR bmi OR diabetes OR ncd* OR menu*
S4 TX student* OR universit* OR "higher education" OR college* OR post-secondar* OR postsecondar* OR campus* OR institution* OR facult* OR graduate OR undergraduate* OR postgraduate* OR "bachelor program*" OR "master program*"
S3 S1 OR S2
S2 (TX dining* OR cafeteria* OR restaurant* OR canteen* OR kitchen OR kitchens OR cuisine OR cuisines OR cater* OR vending OR kiosk* OR "snack bar*" OR snackbar* OR cafe* OR "lunch room*" OR lunchroom* OR foodservice*) OR (TX ( (meals N2 (service* OR system* OR purchas* OR competitiv*)) ) OR TX ( (meal N2 (service* OR system* OR purchas* OR competitiv*)) ) OR TX ( (food N2 (service* OR system* OR purchas* OR competitiv*)) ) OR TX ( (foods N2 (service* OR system* OR purchas* OR competitiv*)) ))

S1 DE "Dining Facilities" OR DE "Breakfast Programs" OR DE "Food Service" OR DE "Lunch Programs"

##### 2.4. Cochrane Library

ID Search
#1 MeSH descriptor: [Food Services] this term only
#2 MeSH descriptor: [Menu Planning] this term only
#3 MeSH descriptor: [Restaurants] this term only
#4 ((food or foods or meal or meals) NEAR/2 (service* or system* or purchas* or competitiv*)):TI,AB,KW
#5 (dining* or cafeteria* or restaurant* or canteen* or kitchen? or kitchens or cuisine or cuisines or "out of home" or cater* or vending or kiosk* or snack bar* or snackbar* or cafe* or lunch room* or lunchroom* or foodservice*):TI,AB,KW
#6 #1 OR #2 OR #3 Or #4 OR #5
#7 MeSH descriptor: [Students] this term only
#8 (student* or universit* or "higher education" or college* or post-secondar* or postsecondar* or campus* or institution* or facult* or graduate or undergraduate or postgraduate or bachelor program* or master program*):TI,AB,Kw
#9 #7 OR #8
#10 MeSH descriptor: [Diet, Food, and Nutrition] explode all trees
#11 (diet* or eating or nutrition* or meal* or food or foods or lunch or beverage* or drink* or refreshment* or water*):TI,AB,Kw
#12 MeSH descriptor: [Sustainable Development] this term only
#13 MeSH descriptor: [Conservation of Natural Resources] this term only
#14 MeSH descriptor: [Climate] this term only
#15 MeSH descriptor: [Climate Change] this term only
#16 MeSH descriptor: [Global Warming] this term only
#17 MeSH descriptor: [Greenhouse Effect] this term only
#18 MeSH descriptor: [Greenhouse Gases] this term only
#19 MeSH descriptor: [Whole Grains] this term only
#20 MeSH descriptor: [Fruit] this term only
#21 MeSH descriptor: [Vegetables] this term only
#22 MeSH descriptor: [Meat] explode all trees
#23 MeSH descriptor: [Vegetarians] explode all trees
#24 (sustainab* or climate or waste* or "eat lancet" or environmental* or carbon or emission or emissions or greenhouse gas* or GHG or global warming or biodiversit* or bio-diversit* or pollution* or planet* or food loss or left over* or leftover* or trash or single-use or reusable or re-usable or locally produced or locally sourced or seasonal or eco* or whole grain* or whole meal or whole meals or wholemeal* or whole wheat or refined grain* or fruit* or vegetable* or legume* or fish* or seafood* or fibre or fibres or sugar* or fat* or dairy or sodium or salt* or water* or calorie* or kilocalorie* or meat* or plant based or foodscape* or vegetarian* or vegan* or flexitarian* or health* or unhealth* or wellbeing or well being or balanced or nutritious or obesit* or over weight* or overweight* or body weight or bodyweigth* or BMI or diabetes or NCD* or menu*):TI,AB,KW

#25 #10 OR #11 OR #12 OR #13 OR #14 OR #15 OR #16 OR #17 OR #18 OR #19 OR #20 OR #21 OR #22 OR #23 OR #24
#26 #6 AND #9 AND #25

#### 2.5. Scopus

( ( TITLE ( dining* OR cafeteria* OR restaurant* OR canteen* OR kitchen OR kitchens OR cuisine OR cuisines OR cater* OR vending OR kiosk* OR "snack bar" OR "snack bars" OR snackbar* OR cafe* OR "lunch room*" OR lunchroom* OR foodservice* OR "out of home" ) ) OR ( ( TITLE ( ( meals W/2 ( service* OR system* OR purchas* OR competitiv* ) ) ) OR TITLE ( ( meal W/2 ( service* OR system* OR purchas* OR competitiv* ) ) ) OR TITLE ( ( food W/2 ( service* OR system* OR purchas* OR competitiv* ) ) ) OR TITLE ( ( foods W/2 ( service* OR system* OR purchas* OR competitiv* ) ) ) ) ) OR ( ( ( TITLE ( ( meals W/2 ( service* OR system* OR purchas* OR competitiv* ) ) ) OR TITLE ( ( meal W/2 ( service* OR system* OR purchas* OR competitiv* ) ) ) OR TITLE ( ( food W/2 ( service* OR system* OR purchas* OR competitiv* ) ) ) OR TITLE ( ( foods W/2 ( service* OR system* OR purchas* OR competitiv* ) ) ) ) ) OR ( TITLE ( dining* OR cafeteria* OR restaurant* OR canteen* OR kitchen OR kitchens OR cuisine OR cuisines OR cater* OR vending OR kiosk* OR "snack bar" OR "snack bars" OR snackbar* OR cafe* OR "lunch room*" OR lunchroom* OR foodservice* OR "out of home" ) ) ) ) AND ( ABS ( ( diet* OR eating OR nutrition* OR lunch OR beverage* OR drink* OR refreshment* OR water* OR sustainab* OR climate OR waste* OR "eat lancet" OR environmental* OR carbon OR emission OR emissions OR "greenhouse gas*" OR ghg OR "global warming" OR biodiversit* OR bio-diversit* OR pollution* OR planet* OR "food loss" OR "left over*" OR leftover* OR trash OR single-use OR reusable OR "re-usable" OR "locally produced" OR "locally sourced" OR seasonal OR eco* OR "whole grain*" OR "whole meal*" OR "whole wheat" OR wholemeal* OR "refined grain*" OR fruit* OR vegetable* OR legume* OR fish* OR seafood* OR fibre OR fibres OR sugar* OR fat* OR dairy OR sodium OR salt* OR water* OR calorie* OR kilocalorie* OR meat* OR "plant based" OR foodscape* OR vegetarian* OR vegan* OR flexitarian* OR health* OR unhealth* OR wellbeing OR "well being" OR balanced OR nutritious OR obesit* OR "over weight*" OR overweight* OR "body weight*" OR bodyweigth* OR bmi OR diabetes OR ncd* OR menu* ) ) ) AND ( ABS ( ( student* OR universit* OR "higher education" OR college* OR post-secondar* OR postsecondar* OR campus* OR institution* OR facult* OR graduate OR undergraduate* OR postgraduate* OR "bachelor program" OR "master program" ) ) )

#### 2.6. Epistemonikos

(title:((title:((student* OR universit* OR "higher education" OR college* OR post-secondar* OR postsecondar* OR campus* OR institution* OR facult* OR graduate OR undergraduate* OR postgraduate* OR "bachelor program" OR "master program")) OR abstract:((student* OR universit* OR "higher education" OR college* OR post-secondar* OR postsecondar* OR campus* OR institution* OR facult* OR graduate OR undergraduate* OR postgraduate* OR "bachelor program" OR "master program"))) AND (title:((diet* OR eating OR nutrition* OR meal* OR food OR foods OR lunch OR beverage* OR drink* OR refreshment* OR water* OR sustainab* OR climate OR waste* OR "eat lancet" OR environmental* OR carbon OR emission OR emissions OR "greenhouse gas*" OR ghg

OR "global warming" OR biodiversit* OR bio-diversit* OR pollution* OR planet* OR "food loss" OR "left over*" OR leftover* OR trash OR single-use OR reusable OR "re-usable" OR "locally produced" OR "locally sourced" OR seasonal OR eco* OR "whole grain*" OR "whole meal*" OR "whole wheat" OR wholemeal* OR "refined grain*" OR fruit* OR vegetable* OR legume* OR fish* OR seafood* OR fibre OR fibres OR sugar* OR fat* OR dairy OR sodium OR salt* OR water* OR calorie* OR kilocalorie* OR meat* OR "plant based" OR foodscape* OR vegetarian* OR vegan* OR flexitarian* OR health* OR unhealth* OR wellbeing OR "well being" OR balanced OR nutritious OR obesit* OR "over weight*" OR overweight* OR "body weight*" OR bodyweigth* OR bmi OR diabetes OR ncd* OR menu*)) OR abstract:((diet* OR eating OR nutrition* OR meal* OR food OR foods OR lunch OR beverage* OR drink* OR refreshment* OR water* OR sustainab* OR climate OR waste* OR "eat lancet" OR environmental* OR carbon OR emission OR emissions OR "greenhouse gas*" OR ghg OR "global warming" OR biodiversit* OR bio-diversit* OR pollution* OR planet* OR "food loss" OR "left over*" OR leftover* OR trash OR single-use OR reusable OR "re-usable" OR "locally produced" OR "locally sourced" OR seasonal OR eco* OR "whole grain*" OR "whole meal*" OR "whole wheat" OR wholemeal* OR "refined grain*" OR fruit* OR vegetable* OR legume* OR fish* OR seafood* OR fibre OR fibres OR sugar* OR fat* OR dairy OR sodium OR salt* OR water* OR calorie* OR kilocalorie* OR meat* OR "plant based" OR foodscape* OR vegetarian* OR vegan* OR flexitarian* OR health* OR unhealth* OR wellbeing OR "well being" OR balanced OR nutritious OR obesit* OR "over weight*" OR overweight* OR "body weight*" OR bodyweigth* OR bmi OR diabetes OR ncd* OR menu*))) AND (title:((((food OR foods OR meal OR meals) AND (service* system* OR purchas* OR competitiv*)) OR ( dining* OR cafeteria* OR restaurant* OR canteen* OR kitchen OR kitchens OR cuisine OR cuisines OR cater* OR vending OR kiosk* OR "snack bar" OR "snack bars" OR snackbar* OR cafe* OR "lunch room*" OR lunchroom* OR foodservice* OR "out of home"))) OR abstract:((((food OR foods OR meal OR meals) AND (service* system* OR purchas* OR competitiv*)) OR ( dining* OR cafeteria* OR restaurant* OR canteen* OR kitchen OR kitchens OR cuisine OR cuisines OR cater* OR vending OR kiosk* OR "snack bar" OR "snack bars" OR snackbar* OR cafe* OR "lunch room*" OR lunchroom* OR foodservice* OR "out of home"))))) OR abstract:((title:((student* OR universit* OR "higher education" OR college* OR post-secondar* OR postsecondar* OR campus* OR institution* OR facult* OR graduate OR undergraduate* OR postgraduate* OR "bachelor program" OR "master program")) OR abstract:((student* OR universit* OR "higher education" OR college* OR post-secondar* OR postsecondar* OR campus* OR institution* OR facult* OR graduate OR undergraduate* OR postgraduate* OR "bachelor program" OR "master program"))) AND (title:((diet* OR eating OR nutrition* OR meal* OR food OR foods OR lunch OR beverage* OR drink* OR refreshment* OR water* OR sustainab* OR climate OR waste* OR "eat lancet" OR environmental* OR carbon OR emission OR emissions OR "greenhouse gas*" OR ghg OR "global warming" OR biodiversit* OR bio-diversit* OR pollution* OR planet* OR "food loss" OR "left over*" OR leftover* OR trash OR single-use OR reusable OR "re-usable" OR "locally produced" OR "locally sourced" OR seasonal OR eco* OR "whole grain*" OR "whole meal*" OR "whole wheat" OR wholemeal* OR "refined grain*" OR fruit* OR vegetable* OR legume* OR fish* OR seafood* OR fibre OR fibres OR sugar* OR fat* OR dairy OR sodium OR salt* OR water* OR calorie* OR kilocalorie* OR meat* OR "plant based" OR foodscape* OR vegetarian* OR vegan* OR flexitarian* OR health* OR unhealth* OR wellbeing OR "well being" OR balanced OR nutritious OR obesit* OR "over weight*" OR overweight* OR "body weight*" OR bodyweigth* OR bmi OR diabetes OR ncd* OR menu*)) OR abstract:((diet* OR eating OR nutrition* OR meal* OR food OR foods OR lunch OR beverage* OR drink* OR refreshment* OR water* OR sustainab* OR climate OR waste* OR "eat lancet" OR environmental* OR carbon OR emission OR emissions OR "greenhouse gas*" OR ghg OR "global warming" OR biodiversit* OR bio-diversit* OR pollution* OR planet* OR "food loss" OR "left over*" OR leftover* OR trash OR single-use OR reusable OR "re-usable" OR "locally produced" OR "locally sourced" OR seasonal OR eco* OR "whole grain*" OR "whole meal*" OR "whole wheat" OR wholemeal* OR "refined grain*" OR fruit* OR vegetable* OR legume* OR fish* OR seafood* OR fibre OR fibres OR sugar* OR fat* OR dairy OR sodium OR salt* OR water* OR calorie* OR kilocalorie* OR meat* OR "plant based" OR foodscape* OR vegetarian* OR vegan* OR flexitarian* OR health* OR unhealth* OR wellbeing OR "well being" OR balanced OR nutritious OR obesit* OR "over weight*" OR overweight* OR "body weight*" OR bodyweigth* OR bmi OR diabetes OR ncd* OR menu*))) AND (title:((((food OR foods OR meal OR meals) AND (service* system* OR purchas* OR competitiv*)) OR ( dining* OR cafeteria* OR restaurant* OR canteen* OR kitchen OR kitchens OR cuisine OR cuisines OR cater* OR vending OR kiosk* OR "snack bar" OR "snack bars" OR snackbar* OR cafe* OR "lunch room*" OR lunchroom* OR foodservice* OR "out of home"))) OR abstract:((((food OR foods OR meal OR meals) AND (service* system* OR purchas* OR competitiv*)) OR ( dining* OR cafeteria* OR restaurant* OR canteen* OR kitchen OR kitchens OR cuisine OR cuisines OR cater* OR vending OR kiosk* OR "snack bar" OR "snack bars" OR snackbar* OR cafe* OR "lunch room*" OR lunchroom* OR foodservice* OR "out of home"))))))

### 3. References used for snowballing searches

Carletto FC, Ferriani LO, Silva DA (2023) Sustainability in food service: A systematic review. Waste Manag Res 41, 285-302

Mingay E, Hart M, Yoong S et al. (2022) The Impact of Modifying Food Service Practices in Secondary Schools Providing a Routine Meal Service on Student's Food Behaviours, Health and Dining Experience: A Systematic Review and Meta-Analysis. Nutrients 14.

Solomou S, Logue J, Reilly S et al. (2023) A systematic review of the association of diet quality with the mental health of university students: implications in health education practice. Health Educ Res 38, 28-68.

Wattick RA, Hagedorn RL, Olfert MD (2018) Relationship between Diet and Mental Health in a Young Adult Appalachian College Population. Nutrients 10.

Plotnikoff RC, Costigan SA, Williams RL et al. (2015) Effectiveness of interventions targeting physical activity, nutrition and healthy weight for university and college students: a systematic review and meta-analysis. Int J Behav Nutr Phys Act 12, 45.

Fonseca LB, Pereira LP, Rodrigues PRM et al. (2021) Food consumption on campus is associated with meal eating patterns among college students. Br J Nutr 126, 53-65.

Sullivan VS, Smeltzer ME, Cox GR et al. (2021) Consumer expectation and responses to environmental sustainability initiatives and their impact in foodservice operations: A systematic review. J Hum Nutr Diet 34, 994-1013.

Kretschmer S, Dehm S (2021) Sustainability Transitions in University Food Service—A Living Lab Approach of Locavore Meal Planning and Procurement. Sustainability 13.

Grech A, Howse E, Boylan S (2020) A scoping review of policies promoting and supporting sustainable food systems in the university setting. Nutrition Journal 19, 97.

Brunner F, Kurz V, Bryngelsson D et al. (2018) Carbon Label at a University Restaurant – Label Implementation and Evaluation. Ecological Economics 146, 658-667.

Visschers VHM, Siegrist M (2015) Does better for the environment mean less tasty? Offering more climate-friendly meals is good for the environment and customer satisfaction. Appetite 95, 475-483.

Thiagarajah K, Getty VM (2013) Impact on Plate Waste of Switching from a Tray to a Trayless Delivery System in a University Dining Hall and Employee Response to the Switch. Journal of the Academy of Nutrition and Dietetics 113, 141-145.

Doherty S, Cawood J, Dooris M (2011) Applying the whole-system settings approach to food within universities. Perspect Public Health 131, 217-224.

Yi S, Kanetkar V, Brauer P (2022) Nudging food service users to choose fruit- and vegetable-rich items: Five field studies. Appetite 173, 105978.

Ringling KM, Marquart LF (2020) Intersection of Diet, Health, and Environment: Land Grant Universities' Role in Creating Platforms for Sustainable Food Systems. Frontiers in Sustainable Food Systems 4.

Lopez V, Teufel J, Gensch C-O (2020) How a Transformation towards Sustainable Community Catering Can Succeed. Sustainability.

### 4. Data extraction sheet

**Study information:**

- Study ID
- Study title
- Publication year

**Study design:**

- Study type (e.g. Impact study, Formative research, Process evaluation, Feasibility study)
- Study design (e.g. Quantitative, Qualitative, Mixed-Methods, Other (like conceptual)
- Methodological approach (e.g. Mathematical modelling, Experimental/ Quasi-experimental, Cross-sectional, Quantitative, Conceptual)
- Focus of the study (e.g. Health, Sustainability, Both)
- Main objective of the study

**Setting:**

- Country
- Setting (e.g. University, College, Vocational School)
- Place of intervention (e.g. Canteen/ Cafeteria, Kiosk, Café, vending machine, Campus-wide, In-class)

**Intervention:**

- Was an actual intervention implemented?
- Main objective of the intervention (e.g. promotion of healthy diet, promotion of sustainable diet, food waste reduction, sustainable food services)
- Detailed objective of the study (e.g. Less plastic/Packaging/ other waste, better energy efficiency, better recycling / composting, improve sourcing practise, healthy diets in general, increase fruit & vegetable intake, increased whole grain intake, lower energy intake, reduced sugar intake, reduced sodium intake, reduced fat intake, reduced intake of animal-based foods, reduced carbon footprint diet, reduce water footprint, Less pre consumer food waste, less post-consumer food waste)
- NOURISHING framework category
- Length of the intervention
- Short description

**Outcomes (repeated for each outcome):**

- Outcome category (e.g. diet quality, diet related health outcomes, mental health outcomes, sustainability outcomes, Acceptability, Feasibility, Uptake, Implementation fidelity, Views on potential intervention(s), Economic)
- Description of outcome
- Outcome attributable to measures (yes/no)
- Level on which outcome is assessed (i.e. students, teachers, staff, wider community, general population)
- Affected / studied population
- Number of participants
- Number of plates / food or drink items sold
- Length of follow-up
- Direction of the effect as judged by the study authors
- Narrative summary

### 5. List of excluded studies

We have listed excluded studies that we deemed potentially interesting to readers in this table. These studies were excluded primarily because they closely aligned with our inclusion criteria but not entirely, prompting extensive discussions within our review team during the screening process.

| Study ID | Study titel | Reason for exclusion | Explanation |
| --- | --- | --- | --- |
| **Rojas 2007** | University of British Columbia Food System Project: Towards Sustainable and Secure Campus Food Systems | No intervention studied | Development of a tool ( Life Cycle Assessment) that could be used to make food services more sustainable. However, the tool is not studied or tested |

| | | | |
|---|---|---|---|
| **Schaubroeck 2018** | A pragmatic framework to score and inform about the environmental sustainability and nutritional profile of canteen meals, a case study on a university canteen | No intervention studied | Development of a tool to score canteen meals, that could be used to make food services more sustainable. However, the tool is not studied or tested |
| **Roy 2021** | Effectiveness of price-reduced meals on purchases among university young adults | Wrong focus | The intervention (price reduced meals) does not aim to promote healthy or sustainable diets, but to alleviate food insecurity |
| **Sato 2020** | Efforts to reduce food loss in restaurants and to assess consumer awareness in japan | No intervention studied | No evaluation of a study. Only assesment of the food waste at the moment and suggestion of possibl interventions |
| **Sherry 2022** | Reducing the environmental impact of food service in universities using life cycle assessment | No intervention studied | Development of a tool ( Life Cycle Assesment) that could be used to make food services more sustainable. However, the tool is not studied or tested |
| **Wongprawmas 2022** | Strategies to Promote Healthy Eating Among University Students: A Qualitative Study Using the Nominal Group Technique | No intervention studied | No evaluation of an intervention. Only assessment of the eating behaviour at the moment and suggestion of possible interventions |
| **Wang 2023** | What influences students' food waste behaviour in campus canteens? | No intervention studied | Influences on the intention to reduce waste is studied, no intervention |
| **Deliberador 2021** | Food waste: evidence from a university dining hall in brazil | No intervention studied | Measurement of current food waste, based on this suggestion of measures. |
| **Frank 2020** | "Free food on campus!": Using instructional technology to reduce university food waste and student food insecurity | Wrong setting | It is about food waste reduction at catered events taking place at the university, not the regular food service operation |
| **Freedman 2011** | Point-of-Purchase Nutrition Information Influences Food-Purchasing Behaviors of College Students: A Pilot Study | Wrong setting | Study takes place in a supermarket that happened to be on campus |
| **Holligan 2017** | Design of Nudge-Based Interventions for Increasing Vegetable Intake in Emerging Adults within On-Campus Dining Sites | No intervention studied | This study is about the process of designing a nudge. The nudge itsels is not tested. |
| **Maher 2017** | Experiential learning for engaging nutrition undergraduates with sustainability | Wrong setting | A class that is taking place at university. However there is no connection to the food service |

| | | | |
|---|---|---|---|
| **Mann 2021** | Development of the University Food Environment Assessment (Uni-Food) Tool and Process to Benchmark the Healthiness, Equity, and Environmental Sustainability of University Food Environments | No intervention studied | Development of a tool ( Life Cycle Assesment) that could be used to make food services more sustainable. However, the tool is not studied or tested |
| **Middha 2021** | Pop-up food provisioning as a sustainable third space: reshaping eating practices at an inner urban university | Wrong focus | The study does look at pop-up food provisioning, but from an architecural point of view. No health or sustainability outcomes related to food are evaluated |
| **Migliavada 2021** | A three-year longitudinal study on the use of pre-ordering in a university canteen | Wrong focus | The intervention does not have "healthy or sustainable diet or food services" as its aim. There is one outcome of "amount of food waste", however it is not compared to the non-pre ordering system, therefore there is no evaluation of the intervention in this point |
| **Monroe 2015** | The Green Eating Project: web-based intervention to promote environmentally conscious eating behaviours in US university students | Wrong setting | No connection to the university food service |
| **Painter 2016** | Food waste generation and potential interventions at Rhodes University, South Africa | No intervention studied | No evaluation of an intervention. Only assessment of the food system at the moment and suggestion of possible interventions |
| **Roys 2016** | The Effect of Energy Labelling on Menus and a Social Marketing Campaign on Food-Purchasing Behaviours of University Students | Wrong setting | A restaurant on campus |
| **Skelton 2020** | A Qualitative Investigation of College Student Perceptions of Their Nutrition Environment: Recommendations for Improvement | No intervention studied | Focus group discussion about the food environment at the moment. Only recommendations for interventions are made, no intervention evaluated |
| **Ulfin 2020** | Monitoring the quality of the cooking oil and raw foods used in canteens in Sepuluh Nopember Institute of Technology to achieve health stalls | Wrong focus | The aim of the study is not really to promote healthier food environments. They simply did a few tests to find out wether the cooking oil meets the national standards. |

### 6. Differences between protocol and review

This study is based on a protocol developed and prospectively registered and published online through the Open Science Framework (registration DOI https://doi.org/10.17605/OSF.IO/CM8VA before data were analyzed. In the following, we describe differences between the protocol and the manuscript:

- We report a slight deviation from our study protocol regarding the inclusion criteria for study designs. We planned to include all study designs (RCTs, observational studies, modeling studies, systematic reviews, scoping reviews, conceptual studies, commentaries, editorials, etc.) but deviated from this approach due to the to a very large number of studies identified. Therefore, we decided to include only primary studies and excluded summaries of evidence (systematic, scoping or narrative reviews).
- In the protocol we planned on doing snowball searches for all included studies. Due to feasibility reasons, we did not manage to do this step. We did, however, in the beginning conduct forward and backward searches in relevant reviews and potentially eligible studies known to the authors.
- Further, we deviated from our published protocol by not presenting our results graphically using Harvest Plots or Effect Direction Plots, as anticipated, as we felt our results were adequately depicted using the abovementioned table using the modified NOURISHING framework and EGM.
